# Chronic Kidney Disease in Ecuador: An Epidemiological and Health System Analysis of an Emerging Public Health Crisis

**DOI:** 10.1101/2021.02.19.21252087

**Authors:** Irene Torres, Rachel Sippy, Kevin Louis Bardosh, Ramya Bhargava, Martín Lotto-Batista, Abigail E. Bideaux, Ramon Garcia-Trabanino, Amelia Goldsmith, Sriram S. Narsipur, Anna M. Stewart-Ibarra

**Affiliations:** Fundacion Octaedro, El Zurriago E8-28 y Ave. De los Shyris, Quito 170505, Ecuador; SUNY Upstate Medical University, Syracuse, NY, USA; University of Florida, Gainesville, FL, USA; Center for One Health Research, School of Public Health, University of Washington, Seattle, WA, USA; SUNY Upstate Medical University, Syracuse, NY, USA; SUNY Upstate Medical University, Syracuse, NY, USA; Helmholtz Centre for Infection Research, Department of Epidemiology, Brunswick, Germany; Centro de Hemodiálisis, San Salvador, El Salvador; Emergency Social Fund for Health, Tierra Blanca, El Salvador; Johns Hopkins University Bloomberg School of Public Health, Baltimore, MD, USA; Inter-American Institute for Global Change Research (IAI), Montevideo, Uruguay; Department of Medicine; SUNY Upstate Medical University, Syracuse, NY, USA

## Abstract

**Background:** The absence of a chronic kidney disease (CKD) registry in Ecuador makes it difficult to assess the burden of disease, but there is an anticipated increase in the incidence of end-stage kidney disease along with increasing diabetes, hypertension and population age. From 2008, augmented funding for renal replacement treatment expanded dialysis clinics and patient coverage.

**Methods:** We conducted 73 in-depth interviews with healthcare providers in eight provinces. Findings were analyzed using qualitative methods and triangulated with quantitative data on patients with CKD diagnoses from six national-level databases between 2015 and 2018. We also reviewed grey and scientific literature on CKD and health systems in Ecuador.

**Results:** Datasets show a total of 17 484 dialysis patients in 2018, or 567 patients per million population (pmp), with an annual cost exceeding 11% of Ecuador’s public health budget. Each year, there were 139—162 pmp new dialysis patients, while doctors report waiting lists. The number of patients on peritoneal dialysis was stable; those on hemodialysis increased over time. Only 13 of 24 provinces have dialysis services, and nephrologists are in major cities, which limits access, delays medical attention, and adds a travel burden on patients. Prevention and screening programs are scarce, while hospitalization is an important reality of CKD patients.

**Conclusion:** CKD is an emerging public health crisis that has increased dramatically over the last decade in Ecuador and is expected to continue, making coverage for all patients impossible and the current structure, unsustainable. A patient registry would permit to estimate the demand and progression of patients with consideration for comorbidities, requirements and costs, and mortality, and identify where to focus prevention efforts. Health policy should clearly state CKD definitions and required patient data, including cause, disease stage and follow-up statistics. Organized monitoring of patients would benefit from improvements in patient referral.

## Introduction

Chronic kidney disease (CKD), defined as “abnormalities of kidney structure or function, present for >3 months, with implications for health” (1), is considered a globally important non-communicable disease (NCD). In 2017, over 9% of the global population, approximately 700 million people, were estimated to suffer from CKD, making it the twelfth leading cause of death (2); another study estimated worldwide prevalence at 11—13% in 2016, making CKD more common than diabetes (3). In Andean Latin America, CKD was the fifth leading cause of death in 2017 (4); in Ecuador, it was estimated as the fourth leading cause of death and fifth leading cause of premature mortality (5).

CKD is a huge burden to health systems; although end-stage kidney disease (ESKD) patients comprise only 0.1—0.2% of developed countries’ population, they require 2—3% of healthcare spending (6). In low-and middle-income countries, it is difficult to assess the exact burden of CKD, but there is an anticipated increase in the incidence of ESKD along with diabetes, hypertension and population age (7,8). The estimated prevalence of ESKD in Latin American is 660 pmp (9). Also, shortage of nephrologists is not uncommon (10). Most preventable deaths from ESKD patients lacking access to renal replacement therapy (RRT) –dialysis or transplantation– occur in low and middle-income countries (11), affecting population health, public health budgets and livelihoods. Ecuador had a population of 16.8 million in 2017, and there were 5739 estimated CKD deaths and 1.2 million estimated prevalent CKD cases (4).

In 2008, Ecuador’s government recognized CKD as a ‘catastrophic illness’ and increased funding for RRT which expanded dialysis clinics and patient coverage. This resulted in better reporting and capture of ESKD but the lack of a systematic registry in accordance with international standards (12) makes it impossible to assess true patterns of CKD epidemiology. Assessing the true incidence and prevalence would help determine resource allocation for management of CKD, reduce the burden of ESKD, and reduce lost productivity. According to the Ministry of Public Health (MSP), an estimated 33 000 people in 2015 were at CKD Stage 5 in Ecuador and 45% of all CKD patients in Stages 4 and 5 (roughly 30 000 people), were expected to die due to unavailability of RRT (5). Research in neighboring Peru reported a similar effect on CKD mortality from insufficient access to dialysis in parts of the country (13).

The structure, policies and functioning of national health systems impact CKD outcomes. Ecuador’s health sector is highly fragmented (14). 41.6% of the population (approximately 7 million) had public health insurance coverage in 2019 (15) through the Ecuadorian Institute of Social Security (IESS in Spanish) and Farmer’s Social Security (SSC in Spanish). An estimated 41.4% of total health spending in Ecuador is out-of-pocket (16), with primary-level care being largely provided by newly-graduated medical professionals as part of the MSP’s one-year compulsory service program.

Access to medical care in Ecuador continues to be a challenge for many, especially in rural areas. Although Ecuador is classified as an “upper middle-income country” by the World Bank (17), 25% of the population live below the poverty line (18), which is defined as a monthly household income of US$84.82 per person. The extensive economic costs of CKD can push entire families into poverty (19), while poverty negatively impacts CKD via poor diet, hazardous occupational conditions, psychosocial stress and sub-optimal access to healthcare (20). Lack of adherence to therapy in CKD negatively impacts quality of care and life (21,22). Poor access to RRT increases mortality in advanced CKD. Limited screening and surveillance, delayed medical care, inadequate training of medical practitioners, and the lack of a national registry limits opportunities for prevention, early detection and management (23).

Public health implications of CKD have been explored for limitations and challenges to the nephrology workforce (24), physician perspectives on the burden and possible causes of CKD (25) or on care planning for patients (26), and the need for improving awareness and screening for prevention (27). Addressing risk factors such as overweight and obesity through integrated or community-based interventions has also gained attention in the past years (28–31), as has the need to prevent the rising prevalence of chronic kidney disease of unknown origin (CKDu, or Mesoamerican nephropathy) in Latin America (32). Studies on CKD patients in Ecuador are limited; the most updated data are from 2014, and are unofficial, relying on practitioners’ estimates collected through a survey (33), which is insufficient to determine health system needs. Reports on outstanding government debt with dialysis providers in recent times point towards financial pressure of ESKD likely escalating in the country (34–36).

We undertook an exploratory, integrated sociologic and epidemiologic study of CKD in Ecuador with the following aims: 1) Estimate the burden of CKD by analyzing databases and other publicly available epidemiologic information; 2) Understand mechanisms of healthcare delivery and its effect on CKD; and 3) Explore the perceptions and experiences of CKD among nephrologists and other physicians.

## Methods

Our study combined qualitative data from healthcare provider’s interviews and quantitative national-level CKD patient data with a review of grey and scientific literature published on CKD and health systems in Ecuador from the MSP and other sources. In this manuscript, we present results in an integrated narrative.

### Ethics

Ethics committee approval (CEU-084) was obtained from the Universidad Internacional del Ecuador; the Institutional Review Board (IRB) of the State University of New York-Upstate Medical University deemed the study protocol exempt. All interviewees were adults (18 years of age or older) and they provided verbal consent for the interview to be recorded. No personal identifying information was recorded. The epidemiological data used in this analysis were anonymized with a unique identifier to allow for duplicate entries to be removed.

### Epidemiological data & analysis

Several national-level datasets were collected for analysis in this paper. There were five datasets from the employee public insurance systems in Ecuador (IESS, SSC, Armed Forces Institute of Social Security, ISSFA, and the Police Force Institute of Social Security, ISSPOL), including retired and dependent persons in these systems, for 2015—2018. These data included all hemodialysis visits at IESS facilities (IESS Hemodialysis), all dialysis procedures (from any facility) billed to the insurance system (IESS Procedures), all visits to an IESS hospital with an 10th International Classification of Disease (ICD-10) code N18 diagnosis (IESS Hospital), all diagnoses of ICD-10 code N18 billed to the insurance system (IESS Diagnosis), and all external (non-IESS facility) consultation or emergency room visits billed to the IESS system with an ICD-10 code N18 diagnosis (IESS Externals). Another dataset from the public health care system in Ecuador included all patients with an N18 diagnosis who used MSP services from 2014—2018. All data were anonymized.

Information on dialysis patients in the IESS system was compiled from the IESS Hemodialysis and IESS Procedures datasets. Individual identification numbers, which were previously anonymized, were compared to eliminate duplicates between datasets and from repeated visits. Summary characteristics were calculated by year at the national level. New patients in the dialysis system were calculated by matching patient identification numbers across years then counting unique patient numbers each year. To project the number of dialysis patients in Ecuador in 2023, IESS and MSP data from 2018, and IESS data from 2015—2017 were used to estimate total (IESS and MSP) dialysis patients for 2015—2018, then total 2023 patients were estimated using dampened Holt’s method (37).

Total IESS patients with a CKD diagnosis were estimated by combining the IESS Hospital, IESS Diagnosis, and IESS Externals datasets. Individual identification numbers were compared to eliminate duplicates between datasets and from repeated visits. Demographic characteristics were calculated by year. This list was cross-checked with the list of dialysis patients in the IESS system to determine the number of individuals with CKD on dialysis in the IESS system. Total MSP patients with a CKD diagnosis were obtained from the MSP dataset. Demographic characteristics were calculated for each year. The IESS Hospitalized dataset was used to assess the number and types of comorbidities among patients with a CKD diagnosis. These include patients with CKD who were hospitalized for any reason. Summary characteristics were calculated by year. The 5 most common comorbidity ICD-10 codes were determined.

Dialysis type was only available for IESS dialysis patients. To estimate the number of patients on hemodialysis and peritoneal dialysis in the MSP system, the proportion of hemodialysis and peritoneal patients in the IESS system were applied to the MSP dialysis patients.

For the IESS system, dialysis patient travel burden was determined using the IESS Diagnosis and IESS Procedures datasets. The location of the clinic where the patient received their initial referral for dialysis was assumed to be within their home province. For each dialysis visit in 2015-2018, the location of the dialysis clinic was compared to the home province and was categorized as the same province, a neighboring (border-sharing) province, or from further away. For the MSP system, patient residence data was available; the location of residence was compared to the location of dialysis clinics during all dialysis visits in 2014-2018, to create the same categories.

Information on health clinics and dialysis services were obtained from the 2008 and 2017 Health Resources and Activities Survey (Recursos y Actividades de Salud, in Spanish) conducted by INEC (https://www.ecuadorencifras.gob.ec/actividades-y-recursos-de-salud/).

### Qualitative interviews

A convenience sample of 73 healthcare providers was selected. First, through discussions with the Nephrology Society of Ecuador, IESS, and MSP from the 286 nephrologists registered to practice, to include as many geographic locations as possible, in both public and private settings. Second, we added nephrologists and physicians located where there are no nephrologists in places recommended by participants.

We conducted 73 in-depth interviews with healthcare providers in eight provinces. This was divided into two phases. The first, spanning 2 weeks in June 2019, included 34 nephrologists and health administrators in the cities Guayaquil (n=19) and Quito (n=15), which are the largest cities and have the highest number of nephrology clinics and services. The second phase spanned 4 weeks between July and August 2019 and included 39 nephrologists, health administrators and physicians in the provinces of Manabí, Guayas, Los Ríos, Cañar, Esmeraldas, Santo Domingo de Los Tsáchilas, Pichincha and Imbabura. Participant details are in Supplemental Table 5.

Phase 1 interviews were conducted in person by two authors (IT and AMS) in 20 hospitals or clinics in Guayaquil and Quito. Phase 2 interviews were conducted in 24 hospitals or clinics, in person (n=28) or by telephone (n=11) by one author (IT). During interviews, we introduced study objectives and obtained oral consent to record the interview. Our interview template included 11 open-ended questions and probes for this study (See Supplemental Text 1) including: professional background, common causes of kidney disease, knowledge and experience with CKD, characteristics of kidney disease patients, standard diagnosis and treatment strategies, and barriers to early detection, treatment and care.

All interviews were conducted in Spanish, audio-recorded, transcribed and translated into English. IT and KB analyzed them. After transcription, all respondents were identified through a code number to ensure anonymity. Transcripts were subsequently coded using a modified grounded theory approach to develop our thematic categories. Analysis involved open coding in the margins of the transcripts. Once all transcripts had been coded and a list of codes developed, the transcripts were revisited using the selective list of codes. Grouping of codes into concepts and then categories relied on memos and sorting. The large sample size supports a reasonable level of saturation.

### Patient and public involvement

This paper does not report a clinical study, so no patients were involved. While the public was not involved in the development and design of the study, we will draft a report of results in Spanish for the Ministry of Public Health of Ecuador and study participants, which we will also disseminate among specialized and other media, and public health professional and advocacy organizations.

## Results

## 1. Increasing incidence and prevalence of ESKD and CKD

### 1.1 Increasing incident/prevalent dialysis patients

Ecuador’s 2008 Constitution recognized CKD as a ‘catastrophic illness’, and from 2012 guarantees medical attention to CKD patients, including dialysis during ESKD (5). According to Ecuador’s Health Activities and Resources Database (38), there were 59,183 dialysis treatments in 2008, which is about 400 chronic patients, assuming 3 treatments per week. Dialysis was unaffordable to most patients before 2012 and ESKD resulted in premature death. The policy change from 2102 meant these patients were now able to seek care:

> *“2008, 2009, more or less, what happened, all the patients who died [before] without diagnosis in their home or any other hospital, became supported by the ministry, and so we had a huge flow of kidney patients that before died without treatment…all those repressed patients suddenly entered the system*.*”* (Quito, Participant 13)

IESS and MSP datasets show a total of 17 484 dialysis patients in 2018 (9641 in IESS and 7843 in MSP), or 567 patients per million population (pmp). Summary statistics for IESS patients are in Supplemental Table 1; detailed data on MSP dialysis patients were not available. Number of patients receiving dialysis annually within the IESS system from 2015-2018 are shown in Figure 1; during this period, the total number of patients increased by more than 2600. Each year, there were 2311—2569 new dialysis patients (*i*.*e*. patients who had not been receiving dialysis the previous year), or 139—162 pmp, likely indicating a high mortality rate among this patient group. The number of patients on peritoneal dialysis was stable (from 350 to 357 patients), while those on hemodialysis increased over the 4 years (from 6663 to 9284 patients). If this annual rate of increase continues, the number of CKD patients needing dialysis by 2023 could reach at least 21 365 (95% confidence interval: 18 255—24 474) or 1170 pmp.

Increased dialysis access has not solved the “silent epidemic” of CKD; most interviewees viewed kidney disease as an emerging public health crisis that increased dramatically over the last decade and is expected to continue: *“the [number of patients] is growing abysmally, just to give you an example. In 2009, there were only 4 dialysis centers in Guayaquil. Now…I counted 16 last year”* (Guayaquil, Participant 3). The number of health facilities offering hemodialysis has grown by 420%, from 19 in 2008 to 76 in 2017 (38,39)(Figure 5), as has the number of hemodialysis machines. Whereas dialysis centers previously had few (3—5) machines, interviewees reported that now many have upwards of 30 each, and most shifts were full. The mean number of machines per facility is 16 (39), and the medical directors who were interviewed reported patient waiting lists.

MSP and IESS databases show increasing CKD diagnoses (Figures 2 & 3). Among patients in the IESS system, the number with CKD increased by 99.3% between 2015—2018, from 14 747 to 29 418. Summary statistics for these patients are in Supplemental Table 2. In the MSP system, the number with CKD increased by 107.3% between 2014—2018, from 14 525 in 2014 to 30 117 in 2018. Summary statistics for these patients are in Supplemental Table 3.

### 1.2. CKD and comorbidities

Interviewees reported that the rise of CKD was linked to the parallel rise in diabetes and hypertension; interviewees estimated that 60% to 90% of all CKD cases in Ecuador were caused by these two NCDs. Causes of CKD mentioned by interviewed nephrologists centered heavily on a syndemic relationship between kidney disease, diabetes, hypertension, obesity, socio-economic status and environmental factors, such as diet (especially salt intake), water quality (brackish, with sediment), chemical exposure and genetic factors. Primarily, the rise in CKD was seen as a result of the rise in other NCDs: *“Go to my [dialysis] room at some point, in one shift, 100% are diabetics”* (Guayaquil, Participant 5). Many complications are triggered by kidney damage, and nephrologists often saw diabetic and hypertensive patients who were referred to them after evaluation by cardiologists, endocrinologists, internal medicine specialists or emergency physicians.

> *“There are prevention programs in the ministry, early care of diabetes, hypertension, obesity…but not chronic kidney disease. Why? Because within the five or seven causes [of death], in the society [of nephrologists], and PAHO and WHO, chronic kidney disease is not the main cause. Rather it is presented as a consequence [of other conditions like diabetes and hypertension]. [But] now we know that it is not only a consequence…”* (Guayaquil, Participant 17).

Comorbidities were common among hospitalized IESS CKD patients. The number of CKD patients with comorbidities are in Supplemental Figure 1. In 2018, 22.1% had no comorbidities, 15.0% had one comorbidity, and 62.9% had two or more comorbidities. The most common comorbidities among patients in 2018 were primary hypertension (7.1%), type-II or type-I diabetes (3.4% each), other urinary tract disorders (2.7%), heart failure (1.7%), and bacterial pneumonia (1.0%) (Figure 4).

## 2. Public health, hospitalization and dialysis service expenditures

With the increasing number of dialysis patients, interviewees lamented the current financial burden of dialysis, and many had dire predictions of the fiscal pressure of these costs in the immediate present and future: *“[dialysis] is a black [fiscal] hole, you put money in it, the money goes, the money goes, the money goes”* (Guayaquil, Participant 5). This was reflected in a dominant belief that the exclusive focus on dialysis has done little to address the risk factors and causes of late-stage CKD. These costs are largely borne by social security schemes that operate under the IESS and MSP, and to a lesser extent, private insurance companies. According to our analysis (Supplemental Table 4), at an annual cost of US$17 472/patient for hemodialysis and US$14 940/patient for peritoneal dialysis, 11.04% of Ecuador’s US$2.665 billion public health budget (40) was spent on dialysis alone in 2018.

Because dialysis machines are imported, some nephrologists interviewed believed that spending limits needed to be better defined and younger patients prioritized for dialysis and transplants. Diabetic patients were also viewed as a transplant priority, because of their high treatment cost; CKD expenses include frequent and long-term hospitalizations for patients on hemodialysis. Many clinicians felt that if unaddressed, CKD poses a major fiscal challenge to the health system:

> *“…we do not have the money to treat all these people, and the budget that one disease eats up, they are going to take away from another… the Ecuadorian state cannot afford not to treat a patient with chronic kidney disease… the [total health] budget is going to become so reduced [due to this], you have no idea”* (Quito, Participant 7).

Hospitalization is an important reality for CKD patients. Among patients in the IESS system, we found that 43.2-48.6% of those with CKD were hospitalized for at least one day annually. In 2018, we found that 3,427 CKD patients were hospitalized, for an average duration of 8.4 days (Table 1).

**Table 1:**
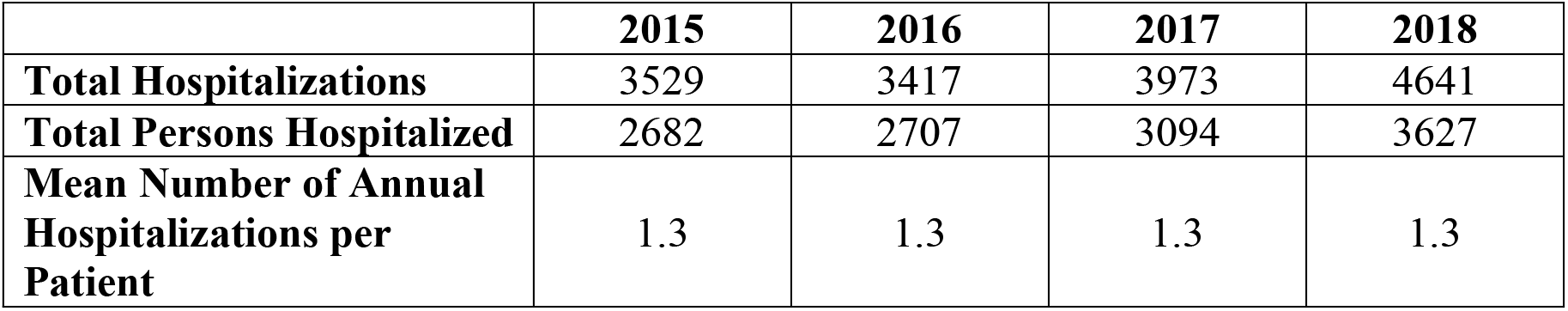
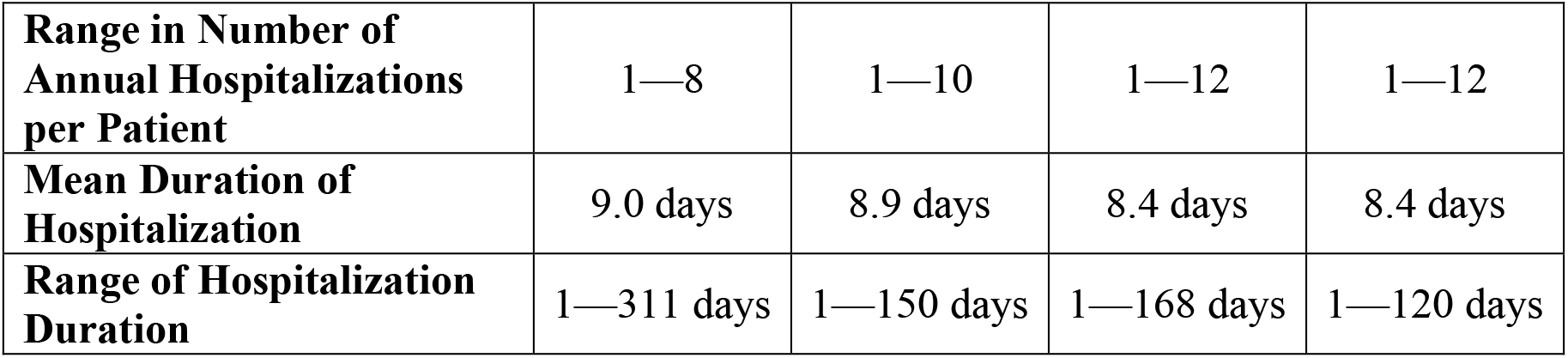
Hospitalization of IESS-Associated Patients with CKD

## 3. Nephrologists, access to treatment and the problem with dialysis

### 3.1. Access to nephrologists and dialysis services

The crisis of CKD means that the primary, secondary and tertiary care networks are overwhelmed with patients seeking dialysis, as reported by our interviewees. Government support for dialysis encouraged proliferation of dialysis centers and access to treatment has increased in major cities (Guayaquil, Cuenca, Quito) as well as in many provincial capitals. While this addressed some access barriers, interviewees explained that dialysis centers remain concentrated in urban areas, with long delays to see nephrologists. In 2019, there were 286 registered nephrologists in Ecuador for a population of 17.2 million people that year, or 16.6 nephrologists pmp. Nephrology is a relatively new speciality in Ecuador and most interviewees reported having studied abroad (Supplemental Table 5).

Quevedo and Esmeraldas, cities of approximately 210 000 people, did not have a nephrologist in the public system during our study period. Some nephrologists we interviewed emphasized the need for nephrologists to be based in “the provinces” to ensure that medium-sized cities are sufficiently covered – otherwise the financial burden of travel for patients can be overwhelming: *“In the rural areas, there is not a single dialysis unit*.*”* (Quito, Participant 13). As of 2017, there were dialysis centers in only 13 of 24 provinces(39). A map showing the national distribution of facilities is shown in Figure 5. The inter-provincial travel necessary for IESS and MSP dialysis patients to access dialysis services is in Supplemental Figures 2 and 3, respectively.

Interviewees explained that healthcare for ESKD patients in rural, peripheral and small cities may not begin at primary care level (Supplemental Figure 4). Patients commonly presented directly to emergency services at local hospitals that lacked a nephrologist or dialysis services, and may die in emergency or intensive care if there are no openings in a dialysis clinic.

In our interviews, nephrologists insisted on the need for individualized care and not a standardized approach: *“human beings cannot be standardized…we think differently, here dialysis is according to what we see is the risk profile of the patient”* (Quito, Participant 13). The cost of dialysis includes treatment and a support package. In private clinics, patients also have access to monthly nutritionist, psychologists, social workers and other medical specialists such as vascular surgeons. By contrast, such care appears to be infrequently available at many MSP and IESS facilities. Most nephrologists, in our interviews, reported low capacity in both private and public systems, with patients not being followed up as indicated in the new CKD guidelines for non-dialysis patients (5).

### 3.2 Life expectancy and the social determinants of dialysis treatment

Mortality in ESKD can be improved with appropriate CKD management. In our interviews, the low life expectancy of many patients was blamed on the asymptomatic presentation of early-stage CKD, making it difficult for patients or primary doctors to detect it: *“so many patients arrive…when 90% of kidney is damaged”* (Quito, Participant 3). Interviewees reported that the lack of symptoms also contributes to a series of problems in patient referral and health screening behavior: people do not accept their diagnosis, especially in the early stages, seek alternative therapies, and as much as 20% may be lost to follow-up until the very late stages. Some nephrologists stressed that patients have a great deal of fear around initiating dialysis:

> *“They arrive with a lot of fear, a lot of disinformation…the patient goes through limbo for months sometimes. We just have a patient who was admitted, who left [X] hospital and has spent two months without dialysis…Why? He is terrified of dialysis because he had a bad hospital experience [in the past]”* (Quito, Participant 13).

Interviewees described the strong psychological and mental health dimension to CKD (20,21). They found that their patients frequently viewed dialysis as a “death sentence” and treatment strained the whole family emotionally and financially. Tragically, some patients are abandoned by their families and are depressed; as one clinician stated: *“those who do not accept the disease are the fastest to go*.*”*

Interviewees identified barriers to health care for CKD patients including the lack of access to health insurance, three times weekly travel to dialysis centers (most providers do not cover travel), specific bureaucratic and administrative issues (e.g. referral to a nephrologist is only valid for 2 months and it can take longer for patients to book an appointment within this window) and the overarching challenge of poverty and low socio-economic status:

> *“people are poor here, they do not have resources…you see the emergency room is full the first days of the month and at end of the month because they have money for the [bus] ticket… sometimes we do not have 100% stock of the medicine required…”* (Guayaquil, Participant 17).

### 3.3 Dialysis treatment guidelines

To address the need for standards of care in CKD, in 2018 the MSP published a set of clinical practice guidelines of CKD, which are described in the publication as “general recommendations” based on international guidelines and scientific evidence by public and private specialists. Although the new guidelines do not explicitly discuss this, nephrologists interviewed emphasized widespread preference for hemodialysis (calculated at 90% of total) rather than peritoneal dialysis. Regionally, the average ratio is 6.6 hemodialysis per peritoneal dialysis patient; in 2018 in Ecuador, we estimated that the ratio was 26.0 (derived from IESS system data).

The focus on hemodialysis means that other modalities of RRT such as peritoneal dialysis and transplants are not encouraged. Many clinicians recommended encouraging peritoneal dialysis (which is often used for children), including using it as a front-line treatment for all patients. However, it is not standard practice for patients to be informed of this option: *“the majority of patients who come to hemo[dialysis] are almost not aware of peri[toneal], especially acute emergency cases when they begin [treatment only] after arriving to emergency”* (Guayaquil, Participant 3). In our interviews, many nephrologists stressed that, compared to hemodialysis, peritoneal is less aggressive, can be performed at home and can prolong life expectancy, especially for older patients, those with comorbidities and young children.

Hemodialysis requires patients to visit dialysis centers three times weekly for roughly four hours each visit. Monthly exams are also required, as is a regimen of medications for complications and other chronic conditions. Peritoneal dialysis is performed at home and requires daily dialysis of approximately 6—8 hours overnight or 4—6 exchanges during the daytime. It requires aseptic measures and training, making family involvement crucial (for hemodialysis, family members must arrange transportation and accompany the patient, since patients are often ill post-dialysis). Other factors limiting the uptake of peritoneal dialysis, according to our interview participants, included: medical team preferences, limited numbers of automated machines, and potential risks, including catheter infections or dysfunction, peritonitis, and anemia. Our interviews described reluctance towards peritoneal dialysis, arguing that patients who are not careful can get infections or because they do not have the necessary hygiene conditions at home. Concurrently, nephrologists emphasized that managing peritoneal treatment is not difficult for family members to learn and should be expanded: *“If you know how to handle an Android phone you know how to operate a peritoneal dialysis machine, and everyone has an Android telephone nowadays, at least that access is universalized, so it is possible”* (Quito, Participant 1).

### 3.4. Transplants

Nephrologists reported underuse of transplants and suggested that restructuring the kidney treatment cascade to include transplants could contribute significantly to life expectancy. According to the National Institute of Organ Donation and Transplants (INDOT, Spanish acronym, 2019), in 2018 only one public hospital and one paediatric public hospital were accredited to perform kidney transplants; we found that there were only 235 kidney transplants in 2018 (41), and 226 in 2019 (42). We estimated an annual transplant rate of 2.4% of dialysis patients in Ecuador for 2018. Nephrologists emphasized the need to promote a “culture of donation” in Ecuador, since the number of donors has not increased significantly. According to one study (43), kidney transplants went from 58 in 2007 to 249 in 2018. INDOT reported in 2018 there were 1623 patients in the kidney waiting list, with 420 new patients (206 more than the previous year); 44% of those on the waiting list are people between 30 to 50 years of age. Given the limited organ supply, there is a need to develop stronger policy guidelines for transplant recipients. Many nephrologists believed that diabetic patients and children should be prioritized due to health expense of both patients and positive health outcomes:

> “*the diabetic patient costs 5 times more than any other patient because he has many comorbidities…and the patient who dies the most in dialysis. 70% die within 5 years, due to cardiovascular problems, so that patient is the one who would benefit most with a transplant…”* (Quito, Participant 11)

## 4. Missing pieces: Early diagnosis and prevention

An unintended consequence of the focus on dialysis has been the creation of a “reactive nephrology” and “expensive medicine” that has: *“transform[ed] us into hemodialysis doctors. That is the problem. That is a major problem”* (Guayaquil, Participant 3). Despite discussions about an integrated healthcare model and agreement about the critical need for preventative interventions, the predominant public health focus has been on terminal treatment. One clinician, reflecting the opinion of others, associated the new Ecuadorean clinical guide for primary care to the American “invasive” terminal approach (which costs more), rather than the European emphasis on more diagnostic testing to identify early CKD cases (Guayaquil, Participant 17).

### 4.1. Urine testing

Our interviews revealed that simple, low-cost urine tests are not widely used to detect the early stages of kidney damage. Some viewed the urine test as a “liquid biopsy” that provided the “first alert” to emerging kidney damage and should be a routine part of medical checkups; others emphasized the need to integrate proteinuria, creatinine and microalbuminuria (urinary sediment) tests in primary care. There was a widely shared sentiment that some general practitioners do not request any kidney tests, despite routinely testing blood glucose, cholesterol, and triglycerides. Others emphasized that using urine tests would require educating the physicians to look for protein in the urine, which is a simple test.

Nephrologists also highlighted the need to address physician interpretation of kidney function tests. They considered an over-emphasis on “normal values” in tests as problematic and the need to calculate kidney function as glomerular filtration rate by validated formulae. MSP’s kidney disease guide for evaluating creatinine levels of early kidney disease lacks specificity in some areas, such as how to evaluate early stages and include multiple factors. Depending on age and gender, and comorbidities such as hypertension and diabetes, values need to be interpreted differently.

### 4.2. Involvement of primary care physicians

Nephrologists frequently mentioned inadequate diagnoses and clinical histories from primary care physicians, contributing to the inability of the health system to address kidney disease at primary and secondary levels of care which has “overwhelmed nephrologists” (Guayaquil, Participant 3). The triage system was seen as “broken”, and nephrologists working with rural patients in cities with 76 000—320 000 people mentioned that not even occupational health physicians at private companies would detect CKD. According to several interviewees, in their experience, up to 100% of patients could present to nephrologists as ESKD, having never received nephrology services before, which is reflected in the few staged CKD individuals in statistics of the national institute of statistics and census (INEC) and the lower number of reported CKD patients compared to MSP and IESS data. Strengthening early detection and prevention would alleviate the strain on nephrologists and address the problem of losing patients to follow-up. Little attention is paid to delaying progression, including the need for dialysis, for example by addressing stage 3 or 4 kidney damage or earlier stages. Some viewed this as a direct result of the “lobbying by companies, the hemodialysis companies” (Guayaquil, Participant 5). Whatever the cause, there was a ubiquitous agreement that prevention is nonexistent and that this reflected a systemic problem in the organization of primary healthcare and health policy:

> *“We have completely deformed the health system… Here everything is atomized [put into vertical programs] and that is what makes us have nothing, in the end*…*we cannot prevent [our patients] from advancing to stage 5 [since there is no focus on prevention]*.*”* (Quito, Participant 13)

To many of the nephrologists we interviewed, the main barrier to early detection was lack of clinical knowledge at lower healthcare levels due to gaps in medical education. Kidney function testing by means of a simple blood test is sparse at the level of primary and secondary care despite the presence of significant co-morbidities such as hypertension and anemia. According to our interviewees, there is a need to re-design nephrology training for medical students, interns and general practitioners, so they have the capacity to recognize early CKD.

### 4.3. Community-based prevention

Our interviews highlighted the need for community-based prevention programs. Although individual nephrologists mentioned some local prevention programs, these were rare, not systematic and many lamented the lack of attention and funding to this critical area. Nephrologists trained abroad emphasized the importance of mass media communication; those working with indigenous populations highlighted the need to work in collaboration with local leaders because traditional medicine is highly valued among them. Some recommended that awareness campaigns become integrated with diabetes and hypertension prevention; others, that these efforts are not reduced to specific events but take place throughout the year. While some dialysis companies have considered prevention, nephrologists expressed concern that the private sector could eclipse national programs to reduce the need for dialysis in the first place. This included community-level screening, urine tests, and an annual kidney profile for high-risk groups, especially those with diabetes and hypertension and those working with agrochemicals. This could be based on existing models for maternal and child health:

*“…95% of kidney diseases [can be identified] in the urine test, which costs 50 cents. Why is it not done? We could just do a screening in the schools, in the community…we have a screening that costs 50 cents and that should be used every year at all levels…to screen, screen at an extremely low price. And if we [combine urine tests with] blood pressure tests*… *almost 100% of kidney diagnoses are made*.*”* (Quito, Participant 13)

Nephrologists also reported a lack of accessible, community-level information about kidney disease, contributing to low knowledge among patients and the wider community. Since CKD is only symptomatic in later stages, many patients are “surprised” and “skeptical” when nephrologists talk about the seriousness of the disease: *“[Some] patients on dialysis do not know about their illness or their treatment, not even in a general way… they are totally unaware. So, I think that…prevention, it is failing, it is failing a lot”* (Quito, Participant 5). Another nephrologist reflected:

> *“In Spain [where I trained], it is impressive because people would come and say “I have this, I have this, I have to take this [medication] and this other” and they would almost come with the diagnosis. Here people arrive, “I take a blue pill but I do not know anything about what it is”…it is important to know the basics of their disease, know how to take care of themselves, what to take, what not to take…”* (Quito, Participant 3).

Discussions about prevention also stressed the need for population-level lifestyle changes and education for changing risk behaviors, specifically for diet (reducing salt), drinking more fluids, and less self-medication with NSAID drugs. Some nephrologists associated specific hotspots of CKD with regional diets in high-salt consumption and junk food such as soda; as one stated: *“Look how obesity has increased…it was not like that in our country [before]. There are so many obese children, who eat on the street, fast foods, and that creates a high risk of kidney failure, hypertension, [and being] overweight…”* (Guayaquil, Participant 14).

## Discussion and Conclusion

This paper examines the burden of CKD in Ecuador by analyzing data from its healthcare system and the perspectives of nephrologists and other healthcare providers. We observed that the number of patients diagnosed with CKD and ESKD has increased over time in Ecuador, with many arriving directly to RRT during emergency care before detection in primary care. CKDu has emerged as a major public health concern in populations with similar labor conditions as those experienced in coastal Ecuador and although this is beyond the scope of the current study, future research is needed to determine whether CKDu or Mesoamerican nephropathy is present in the country, as has been documented in Central America and other countries.

Similar to other countries in Latin America (13), healthcare coverage is not available for all patients with CKD in Ecuador. Not all provinces have nephrologists working in the public sector, or at all, and all are in urban areas. Depending on place of residence, patients may have to travel to access dialysis services. Care restricted to urban areas is a major barrier to access and implies costly and lengthy transportation as well as delays in medical attention. Also comparable to the rest of the region (33), peritoneal dialysis is underutilized and represents an opportunity to improve quality of life and life expectancy, and reduce patient financial burden.

This study has several limitations. Our estimate of 567 dialysis patients pmp in Ecuador does not include all possible ESKD patients in the country including undiagnosed or unreported cases, patients on waiting lists for dialysis services, and patients possibly utilizing private healthcare systems. As in other Latin American countries (33), the lack of data on CKD staging and RRT waiting lists limits our assessment of CKD and ESKD. Without a systematic registry, it is not possible to calculate incidence nor fully evaluate CKD epidemiology. The datasets did not have the detailed level of medical records, meaning we had an incomplete picture of each patient’s health. Data on comorbidities was restricted to those in IESS system, and could also be underreported, therefore their burden among CKD patients cannot be quantified more precisely. However, the datasets analyzed here represent the most complete national data available, compiling for the first time data from both the MSP and IESS systems.

This research exposes critical health measures and policy actions for Ecuador. CKD cannot be managed without a national patient registry including morbidity and mortality data. Kidney disease registries permit the estimation of demand and progression of patients with consideration for comorbidities, calculation of costs and health care services, and identification of geographic hotspots in which to focus prevention efforts. Health policy should clearly state CKD definitions and required patient data, including disease stage and necessary follow-up statistics. Together with improvements in referral and counter-referral with considerations for data confidentiality and security, this will facilitate more organized monitoring of patients.

Ecuador has fewer nephrologists pmp than other countries in the region, such as Argentina and Uruguay, with 34.54 and 46.98 nephrologists pmp, respectively (10). Because CKD and demand for RRT are prevalent in all provinces of Ecuador, but there are limited opportunities to specialize in nephrology locally, the government should consider supporting in-country specialization programs as well as redistributing nephrologists or defining a strategy to secure nephrology consultation through the public sector at least in main cities. Improved identification of CKD could be achieved by addressing possible gaps in medical education by re-training physicians at the primary level and changing curricular guidelines to focus on prevention, early detection, and delaying progression.

Repeated reports of government nonpayment to dialysis providers show that pressure on the public health sector is already at a breaking point (34–36). This research highlights the need for CKD to be recognized by the healthcare system as a catastrophic illness deserving free-of-cost coverage necessitates that a significant portion of public health expenditure be redirected go to dialysis services, which may soon become unsustainable. Hospitalization costs additionally contribute to these expenditures.

National policymakers should anticipate increased healthcare utilization in the coming years by patients with obesity-related comorbidities such as hypertension and diabetes. The government must create wider policy interventions to improve awareness and knowledge of CKD and related NCDs in the general population. In some countries, conditions related to CKD such as overweight and obesity are being addressed through public health measures, (e.g. food labelling, restrictions on unhealthy food advertising and media campaigns to change consumer behavior) (31). Interventions integrated in development programs (28) and community-based health promotion (29) also show promise, as do strategies using community health workers (30).

## Data Availability

Data availability
Health system data are available upon request from the Ecuadorian Ministry of Public Health and Ecuadorian Institute of Social Security.

## Acknowledgements

We thank Fundacion Octaedro and the Ministry of Public Health for procuring and providing deidentified data and thank all of the people who participated in the study.

