## Supplemental Materials for "Chronic Kidney Disease in Ecuador: An Epidemiological and Health System Analysis of an Emerging Public Health Crisis"

**Supplemental Table 1: IESS Patients on Dialysis, 2015—2018**

Data are provided as numbers or percentages as indicated.

| *Year* | | ***2015*** | ***2016*** | ***2017*** | ***2018*** |
|---|---|---|---|---|---|
| *Total Dialysis Visits* | | 64 129 | 73 553 | 81 171 | 89 565 |
| *Type of Visit* | *Hemodialysis* | 61 265<br>95.5% | 70 238<br>95.5% | 77 807<br>95.9% | 86 007<br>96.0% |
| | *Peritoneal Dialysis* | 2864<br>4.5% | 3315<br>4.5% | 3364<br>4.1% | 3558<br>4.0% |
| *Mean Visits per Patient* | | 9.1 | 9.3 | 9.3 | 9.3 |
| *Patients* | | 7013 | 7922 | 8722 | 9641 |
| *Sex* | *Male* | 4488<br>64.0% | 5701<br>64.0% | 5573<br>63.9% | 6174<br>64.0% |
| | *Female* | 2525<br>36.0% | 2221<br>36.0% | 3 149<br>37.1% | 3467<br>36.0% |
| *Ages* | *0-10* | 1 532<br>21.8% | 1 493<br>18.8% | 1 452<br>16.6% | 1 356<br>14.1% |
| | *11-20* | 74<br>1.1% | 101<br>1.3% | 113<br>1.3% | 115<br>1.2% |
| | *21-30* | 188<br>2.7% | 247<br>3.1% | 266<br>3.0% | 302<br>3.1% |
| | *31-40* | 356<br>5.1% | 392<br>4.9% | 474<br>5.4% | 489<br>5.1% |
| | *41-50* | 563<br>8.0% | 648<br>8.2% | 705<br>8.1% | 810<br>8.4% |
| | *51-60* | 1136<br>16.2% | 1383<br>17.5% | 1513<br>17.3% | 1678<br>17.4% |
| | *61-70* | 1376<br>19.6% | 1661<br>21.0% | 1989<br>22.8% | 2324<br>28.1% |
| | *71-80* | 799<br>11.4% | 992<br>12.5% | 1189<br>13.6% | 1470<br>15.2% |
| | *81+* | 250<br>3.6% | 347<br>4.4% | 395<br>4.5% | 514<br>5.3% |
| | *Missing* | 739<br>10.5% | 658<br>8.3% | 626<br>7.2% | 583<br>6.0% |

**Supplemental Table 2: IESS Patients with CKD, 2015—2018**

Data are provided as numbers or percentages as indicated.

| *Year* | | **2015** | **2016** | **2017** | **2018** |
|---|---|---|---|---|---|
| *Patients* | | 14 757 | 18 917 | 23 926 | 29 418 |
| *Sex* | *Male* | 9609<br>65.1% | 12 247<br>64.7% | 15 666<br>65.5% | 19 451<br>66.1% |
| | *Female* | 5148<br>34.9% | 6670<br>35.3% | 8260<br>34.5% | 9967<br>33.9% |
| *Ages* | *0-10* | 726<br>4.8% | 716<br>3.8% | 696<br>2.9% | 867<br>2.9% |
| | *11-20* | 116<br>2.1% | 152<br>0.8% | 207<br>0.9% | 222<br>0.8% |
| | *21-30* | 303<br>4.2% | 357<br>1.9% | 384<br>1.6% | 469<br>1.6% |
| | *31-40* | 625<br>7.3% | 662<br>3.5% | 817<br>3.4% | 909<br>3.1% |
| | *41-50* | 1073<br>17.4% | 1283<br>6.8% | 1591<br>6.6% | 1868<br>6.3% |
| | *51-60* | 2561<br>24.4% | 3097<br>16.4% | 3868<br>16.2% | 4555<br>15.5% |
| | *61-70* | 3595<br>21.4% | 4824<br>25.5% | 6402<br>26.8% | 7973<br>27.1% |
| | *71-80* | 1864<br>12.6% | 4211<br>22.3% | 5533<br>23.1% | 6941<br>23.6% |
| | *81+* | 743<br>5.0% | 2690<br>14.2% | 3476<br>14.5% | 4400<br>15.0% |
| | *Missing* | 739<br>10.5% | 925<br>4.9% | 952<br>4.0% | 1214<br>4.1% |

**Supplemental Table 3: MSP Patients with CKD, 2014—2018**

Data are provided as numbers or percentages as indicated.

| *Year* | | **2014** | **2015** | **2016** | **2017** | **2018** |
|---|---|---|---|---|---|---|
| *Patients* | | 14 525 | 17 161 | 20 458 | 21 920 | 30 117 |
| *Sex* | *Male* | 7233<br>49.8% | 8735<br>50.9% | 10 507<br>51.4% | 11 207<br>51.1% | 15 784<br>52.4% |
| | *Female* | 7292<br>50.2% | 8425<br>49.1% | 9937<br>48.6% | 10 689<br>48.8% | 14 326<br>47.6% |
| | *Intersex* | 0<br>0.0% | 1<br>0.0% | 14<br>0.1% | 17<br>0.1% | 7<br>0.0% |
| *Ages* | *0-10* | 119<br>0.8% | 136<br>0.8% | 108<br>0.5% | 116<br>0.5% | 206<br>0.7% |
| | *11-20* | 360<br>2.5% | 418<br>2.4% | 452<br>2.2% | 242<br>1.9% | 521<br>1.7% |
| | *21-30* | 761<br>5.2% | 827<br>4.8% | 983<br>4.8% | 1032<br>4.7% | 1266<br>4.2% |
| | *31-40* | 965<br>6.6% | 1143<br>6.7% | 1295<br>6.3% | 1279<br>5.8% | 1581<br>5.2% |
| | *41-50* | 1847<br>12.7% | 2083<br>12.1% | 2292<br>11.2% | 2416<br>11.0% | 3032<br>10.1% |
| | *51-60* | 3192<br>22.0% | 3606<br>21.0% | 4425<br>21.6% | 4582<br>20.9% | 6271<br>20.8% |
| | *61-70* | 3428<br>23.6% | 4066<br>23.8% | 5067<br>24.8% | 5576<br>25.4% | 7646<br>25.4% |
| | *71-80* | 2521<br>17.4% | 3008<br>17.5% | 3665<br>17.9% | 3988<br>18.2% | 5830<br>19.4% |
| | *81+* | 1323<br>9.1% | 1863<br>10.9% | 2146<br>10.5% | 2498<br>11.4% | 3764<br>12.5% |
| | *Missing* | 9<br>0.1% | 0<br>0.0% | 25<br>0.1% | 9<br>0.0% | 0<br>0.0% |

**Supplemental Table 4: Estimated cost of dialysis to the Ecuadorian public health system**

| | *Monthly cost per patient* (44) | *Annual cost per patient* | *Number of patients in 2018* | *Total estimated annual cost in 2018* |
|---|---|---|---|---|
| Hemodialysis | US$1456 | US$17 472 | 16 837 | US$294 176 064 |
| Peritoneal | US$1245 | US$14 940 | 647 | US$9 666 180 |
| TOTAL | | | 17 484 | US$303 842 244 |

### Supplemental Table 5: Characteristics of Interview Participants

| Category | | Total sample (n=73) |
|---|---|---|
| **Province** | Guayas | 23 (31.5%) |
| | Pichincha | 17 (23.3%) |
| | Manabí | 10 (13.7) |
| | Los Ríos | 13 (17.8%) |
| | Imbabura | 4 (5.5%) |
| | Cañar | 4 (5.5%) |
| | Other | 2 (2.7%) |
| **Gender** | Male | 46 (63%) |
| | Female | 27 (37%) |
| **Type of facility** | IESS | 31 (42.5%) |
| | Private | 27 (37%) |
| | MSP | 13 (17.8%) |
| | Non-profit | 2 (2.7%) |
| **Role(s)[1]** | Clinical nephrologist | 60 (41.7%) |
| | University professor | 3 (2.1%) |
| | Transplant program | 2 (1.4%) |
| | Dialysis services | 34 (23.6%) |
| | Medical director | 32 (22.2%) |
| | General physician/internist | 12 (8.3%) |
| | Resident doctor | 1 (0.7%) |
| **Years of experience** | 0-3 | 8 (11%) |
| | 4-10 | 20 (27.4%) |
| | 10+ years | 24 (32.9%) |
| | N/A | 21 (28.8%) |
| **Nephrology training conducted outside Ecuador** | Yes | 27 (45%) |
| | No | 7 (11.7%) |
| | N/A | 26 (43.3%) |
| **Average number of patients seen per week[2]** | 0-20 | 9 (12.3%) |
| | 20-40 | 15 (20.5%) |
| | 40-80 | 16 (21.9%) |
| | 80+ | 14 (19.2) |
| | Not applicable or available | 19 (26%) |

[1]Multiple roles permitted per participant
[2]Monthly number of patients was divided by four when necessary
IESS=Instituto Ecuatoriano de Seguridad Social, MSP=Ministerio de Salud Pública

**Supplemental Figure 1: Comorbidities among IESS Patients with CKD, 2015—2018**

Comorbidity data were available for IESS patients with CKD who had been hospitalized in each year. The total number of comorbidities for each patient are in the plot.

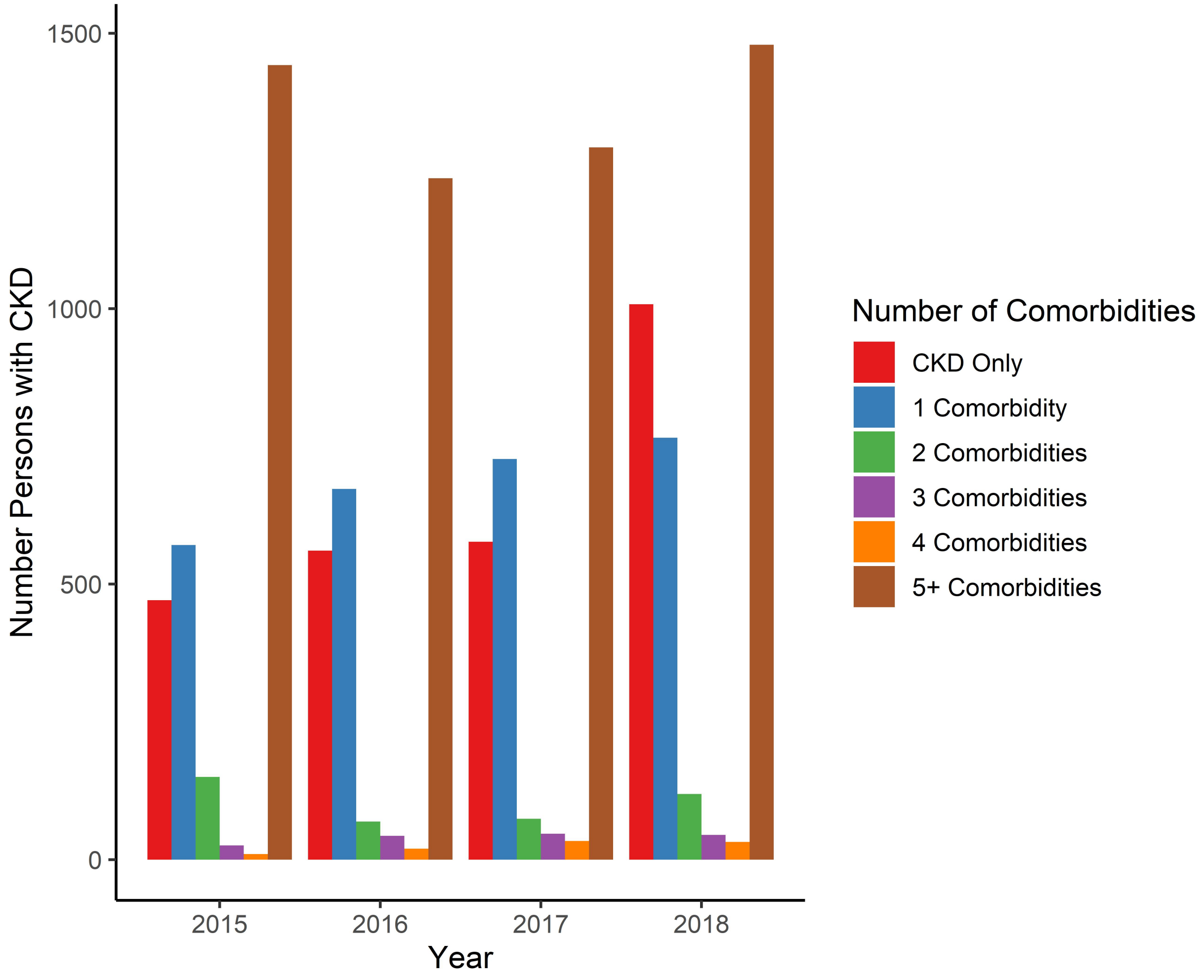

**Supplemental Figure 2: Patient Travel to IESS Dialysis Clinics, 2015-2018**

Data include all dialysis service visits from 2015-2018 for patients in the IESS system, which covers 17 provinces out of 24. The location of their initial referral was compared to visits for dialysis services and whether dialysis was provided in the same province as the referral (green), a neighboring province (blue), or from farther away (red). Patients from Los Ríos, Chimborazo, and Loja had to travel most frequently to another province for dialysis.

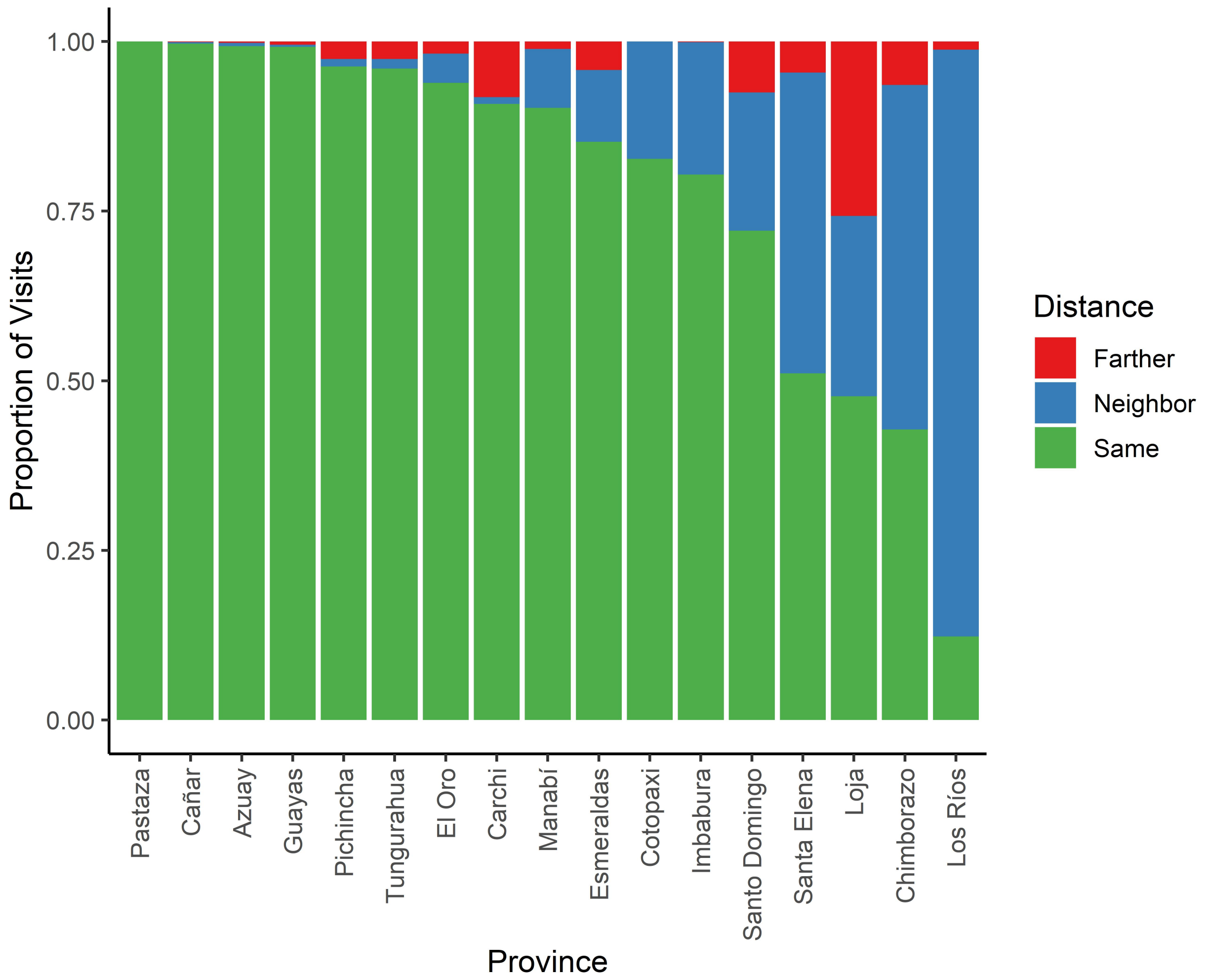

**Supplemental Figure 3: Patient Travel for MSP Services, 2014-2018**

Data include all CKD-related visits from 2014-2018 for patients in the MSP system, which covers 23 provinces out of 24 (Galápagos province lacks dialysis services). The location of patient residence was compared to visits for CKD-related visits and whether these visits were the same province as the patient's residence (green), a neighboring province (blue), or from farther away (red). Patients from Bolívar and Carchi in the Andean highlands, and Zamora Chinchipe and Orellana in the Amazon region, had to travel most frequently to another province for CKD-related service.

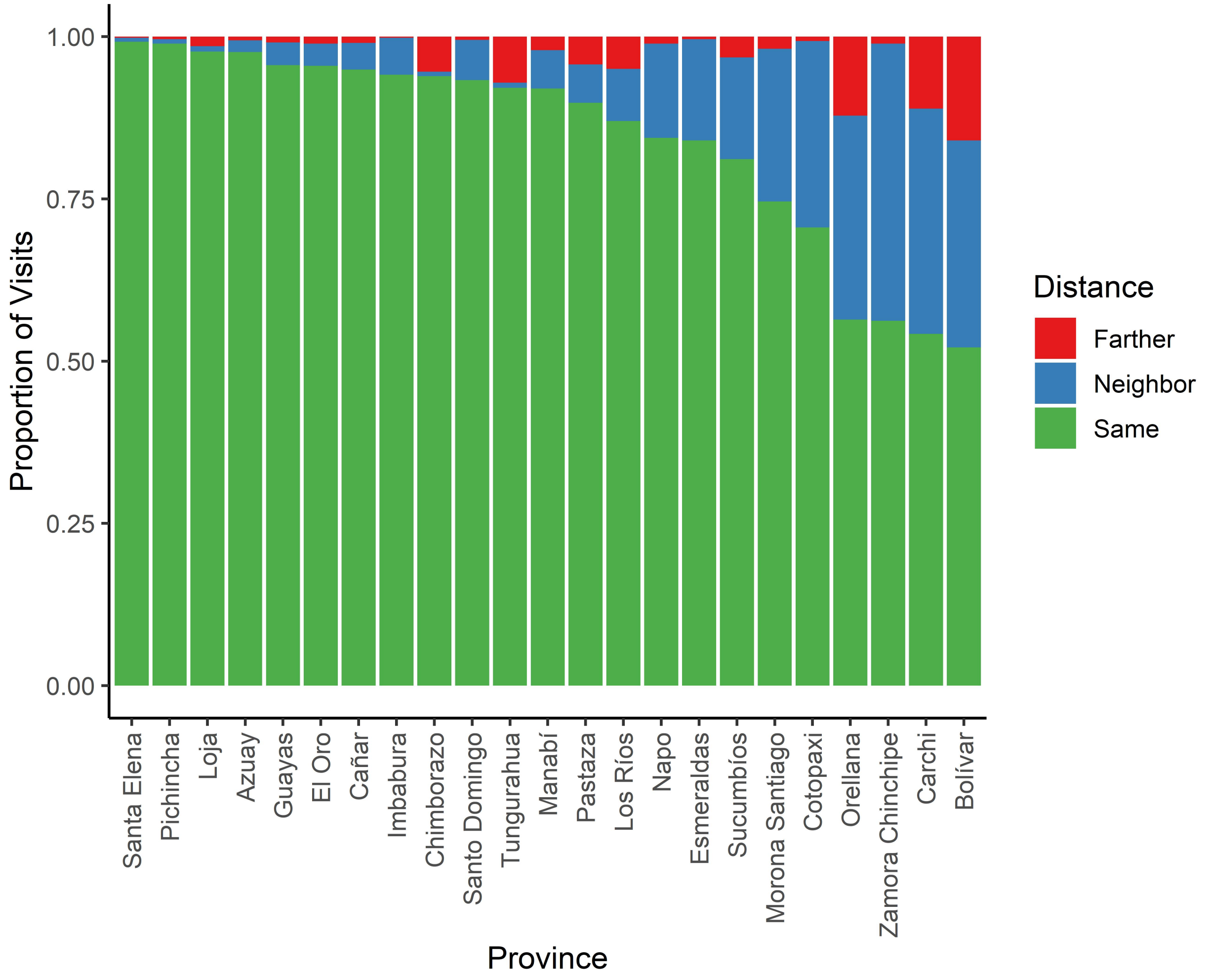

**Supplemental Figure 4: Ecuadorian Health Services Network from the Perspective of an End-Stage CKD Patient**

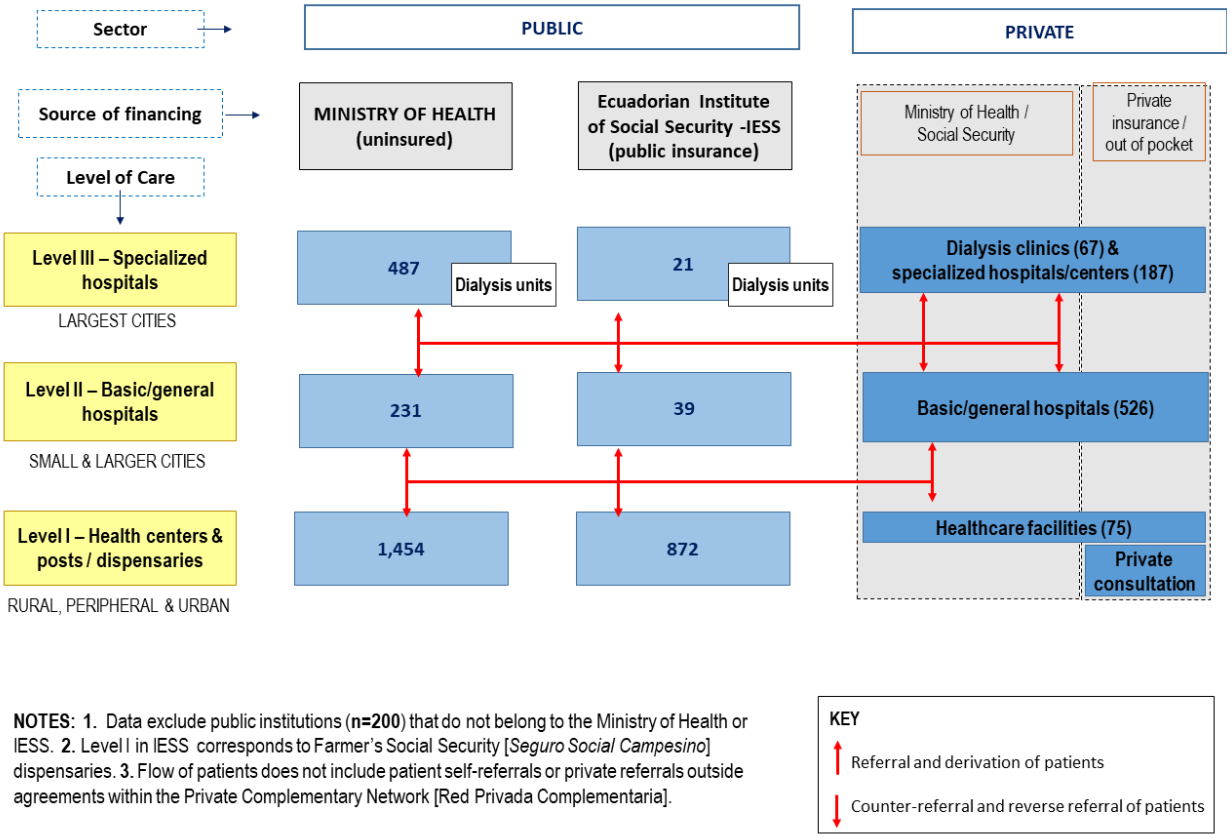


SOURCES: Statistical Registry of Health Resources and Activities (39); Government of Ecuador Official Register Nº 428, Supplement, January 30, 2015; Ministry of Health Technical Guidelines 2014 [*Norma técnica Subsistema de referencia, derivación, contrareferencia, referencia inversa y transferencia del Sistema Nacional de Salud*]; Licensed kidney health specialized centers 2019 [*Centros especializados en salud renal con licencias emitidas*]; Interviews.

### Supplemental Text 1: Key Informant Interview Questions

**1. Can you tell me about your professional background and current position?**

Probe:

- How long have you been in practice?
- How many kidney disease patients do you see a week, on average?
- How many of these patients are on dialysis?
- Describe your place of work?

**2. What are the common causes of kidney disease in your practice?**

Probe:

- Where do most of your kidney disease patients come from (urban, rural, coast, Andes)?
    - Why do you think most of them come from this area(s)?
- Do you think kidney disease has increased in prevalence in your area over the last 10-20 years?
    - If yes, why? Explain.

**3. What is your patient profile?**

Probe:

- How many patients do you see per week on average?
- Do your CKD patients share any common socio-demographic factors, like gender, age, profession, socioeconomic status, place of residence, ethnicity, or something else? Explain.
- What type of patients bears the greatest burden of disease?

**4. What are the common presentations of CKD that you see in your clinic?**

Probe:

- Do your patients with kidney disease have access to prior nephrology care (pre-dialysis) before coming to you?
- Do you receive patients from other providers, or send patients to other providers?

**4. According to your understanding, what are the key factors that increase the risk of CKD?**

**5. Do you think CKD has increased in prevalence over the last 10-20 years in your area?**

Probe:

- Why or why not?
- How significant of a public health issue is CKD in Ecuador?
- Do you think CKD is going to increase more in the future? Why or why not?

**6. According to your understanding, what do your patients and the wider community think are the causes of CKD?**

**7. Can you tell me about your CKD patients and how you treat and care for them?**

Probe

- What other problems have you encountered in attempting to care for CKD patients?
- Do you have many patients that are lost to follow-up? Deaths?

**8. At what stage of disease do you diagnose CKD the most?**

Probe:

- For example, end-stage kidney disease – Needing dialysis
- For example, advanced kidney disease – requiring getting the patient ready for dialysis (CKD stage 4-5)
- For example, early kidney disease (CKD stage 1 to 3)

**9. What barriers to early detection have you experienced with your patients?**

Probe:

- Patient's lack of access to health care
- Lack of lab tests
- Non-availability of a national database for kidney diseases
- Lack of population screening program
- Stigma
- Socio-cultural issues around health seeking behavior

**10. What barriers to treatment and care have you experienced with your CKD patients?**

Probe:

- Patient's lack of access to health care
- Lack of medication
- Poor health seeking behavior
- Socio-cultural issues

**11. Do you believe that there is enough attention being given to CKD in Ecuador?**

Probe:

- Do you feel that CDK is attracting enough attention? Why or why not?
- Are there any health education campaigns focused on kidney disease?
    - If yes, tell me about them. What was good about them and what was not so good?
- What needs to be done to raise the visibility of kidney disease and CKD in particular?
- What resources would help you diagnose and manage kidney disease and CKD better, to have better patient outcomes?
    - Point of care tests (blood and/or urine tests)
    - Greater community awareness?
    - Greater awareness of the problem among other health staff? (referral problems)
    - Community screening programs?
